# Using ICU data to improve the real-time estimation of the effective reproductive number of the COVID-19 epidemic in 9 European countries

**DOI:** 10.1101/2020.04.13.20063388

**Authors:** Samuel Hurtado, David Tinajero

## Abstract

We replicate a recent study by the Imperial College COVID-19 Response Team (Flaxman et al, 2020) that estimates both the effective reproductive number, R_t_, of the current COVID-19 epidemic in 11 European countries, and the impact of different nonpharmaceutical interventions that have been implemented to try to contain the epidemic, including case isolation, the closure of schools and universities, banning of mass gatherings and/or public events, and most recently, widescale social distancing including local and national lockdowns. The main indicator they use for measuring the evolution of the epidemic is the daily number of deaths by COVID-19 in each country, which is a better statistic than the number of identified cases because it doesn’t depend so much on the testing strategy that is in place in each country at each moment in time.

We improve on their estimation by using data from the number of patients in intensive care, which provides two advantages over the number of deaths: first, it can be used to construct a signal with less bias: as the healthcare system of a country reaches saturation, the mortality rate would be expected to increase, which would bias the estimates of R_t_ and of the impact of measures implemented to contain the epidemic; and second, it is a signal with less lag, as the time from onset of symptoms to ICU admission is shorter than the time from onset to death (on average, 7.5 days shorter). The intensive care signal we use is not just the number of people in ICU, as this would also be biased if the healthcare system has reached saturation (in this case, biased downwards, as admissions are no longer possible when all units are in use). Instead, we estimate the daily demand of intensive care, as the sum of two components: the part that is satisfied (new ICU admissions) and the part that is not (which results in excess mortality).

**Thanks to the advantages of this ICU signal in terms of timeliness and bias, we find that most of the countries in the study have already reached R**_**t**_**<1 with 95% confidence (Italy, Spain, Austria, Denmark, France, Norway and Switzerland, but not Belgium or Sweden), whereas the original methodology of Flaxman et al (2020), even with updated data, would only find R**_**t**_**<1 with 95% confidence for Italy and Switzerland**.

## 2. The COVID-19 intensive care signal

We construct an indicator of demand of intensive care by combining the number of deaths by COVID-19 with the number of admissions into intensive care and what is known about the time of evolution of the disease.

Bhatraju et al (2020) describes the distribution of time in intensive care and final result (death or discharge) for 24 patients in the region of Seattle. According to this data, the mortality rate among ICU patients is 50%, with the distribution of daily probabilities of death and discharge that we present in figure 1. It must be recognized, though, that there is great uncertainty around these estimates: Wei-jie Guan et al (2020), using data from 55 ICU patients in several Chinese hospitals until 29^th^ of January of 2020, estimates a mortality rate of 20%, whereas Fei Zhou et al (2020) estimates a mortality rate of 78% using data from 50 ICU patients from two hospitals in Wuhan before 31^st^ January 2020. The aggregation of all of these results would lead an estimated mortality rate of 48%, close to the one reported by Bhatraju et al (2020), which in any case is the one we use because it includes the full distribution of times from ICU admission to death or discharge.

**Figure 1:**
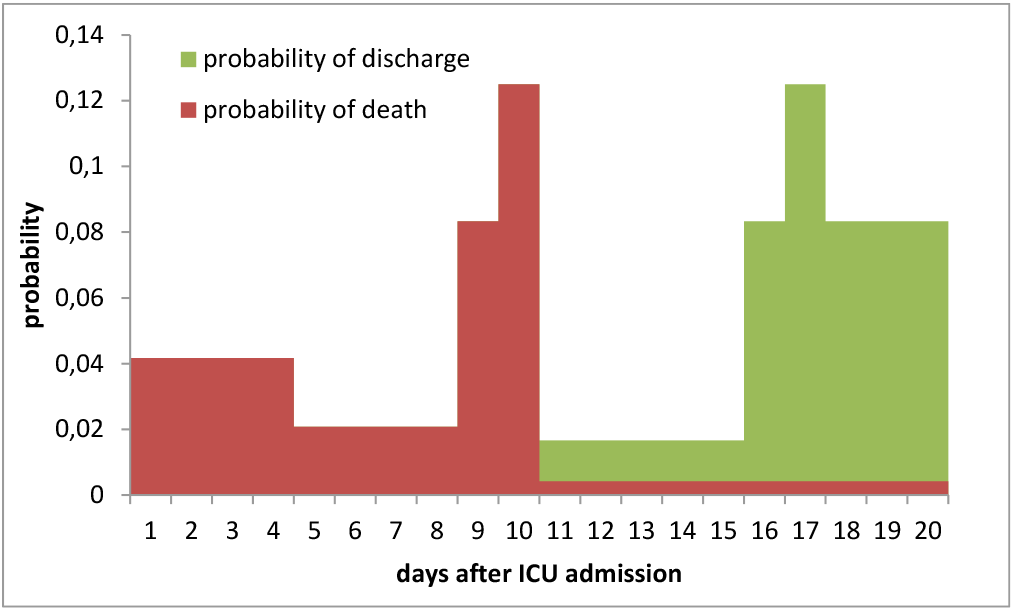
Evolution of ICU patients according to Bhatraju et al (2020) (smoothed)

Using this distribution and the number of COVID-19 patients in intensive care, we can estimate the number of deaths coming from ICU patients, and compare this with the total number of deaths in each country to calculate, through this excess mortality, the demand for intensive care that has not been met. For this we assume that patients in need of intensive care that could not get admission into ICU will die in the following two days. Two facts allow us to confidently make this assumption: first, an extremely high mortality rate is to be expected for this group, as patients that get intensive care already face a 50% mortality rate; and second, we know this happens very fast, because in Spain, whose healthcare system has been saturated during the epidemic (as of today, 10,468 people have received intensive care and 16,353 people have died) average time from onset of symptoms to ICU admission and from onset of symptoms to death is basically identical: 8 vs 9 days, with interquartile ranges of 5-10 and 5-12 respectively^1^. Given this information, it seems like a reasonably conservative assumption.

The estimation is done through a simple inflow-outflow model of the number of people in intensive care (see appendix), using the lag distribution of Bhatraju et al (2020). In the case of Spain, this is done separately by region and then aggregated, as different regional governments publish numbers of ICU patients in terms of prevalence (people in ICU today) or cumulative figures (total number of people that have been in ICU until today).

This signal provides information about the evolution of the epidemic with an approximate lag of 14.5 days: average time from contagion to death (20 days), minus average time from ICU admission to death (7.5 days), plus two days of extra lag because of the assumption about the mortality of patients in need of intensive care but not receiving it. It is therefore a more timely indicator than the number of deaths, which has an approximate lag of 20 days. But it also adequately models the change in that information lag as the epidemic progresses and the healthcare system becomes saturated: when a higher portion of patients requiring critical care is unable to receive it, the time from contagion to death shortens. The mechanical ICU inflow-outflow model also provides information about how saturated the healthcare system is in each country.

Figure 2 presents the results of this estimation of the demand of intensive care in nine European countries: those included in the paper by Flaxman et al (2020), minus United Kingdom and Germany, that don’t publish the necessary information about people in intensive care. Additionally, the data for France only includes deaths in hospitals, as the figures for deaths in retirement homes has been published late and without the necessary detail about date of death. The figures separate the two components (met and unmet demand) and therefore show the different degrees of saturation of the healthcare systems in different countries. According to this, Italy has been able to meet a lower share of the demand for intensive care than other countries, including Spain, probably because the epidemic has been more concentrated (Lombardy represents 52% of the ICU demand in Italy, whereas the region of Madrid represents 36% of the demand in Spain). On the other extreme, results for countries such as Austria, Denmark and Norway show that they have been able to provide intensive care to almost all the COVID-19 patients that required it, in part because the epidemic reached a smaller relative size. The case of France, which has both a large epidemic and high coverage rate, is probably an artefact of the exclusion of deaths in retirement homes in the data.

**Figure 2:**
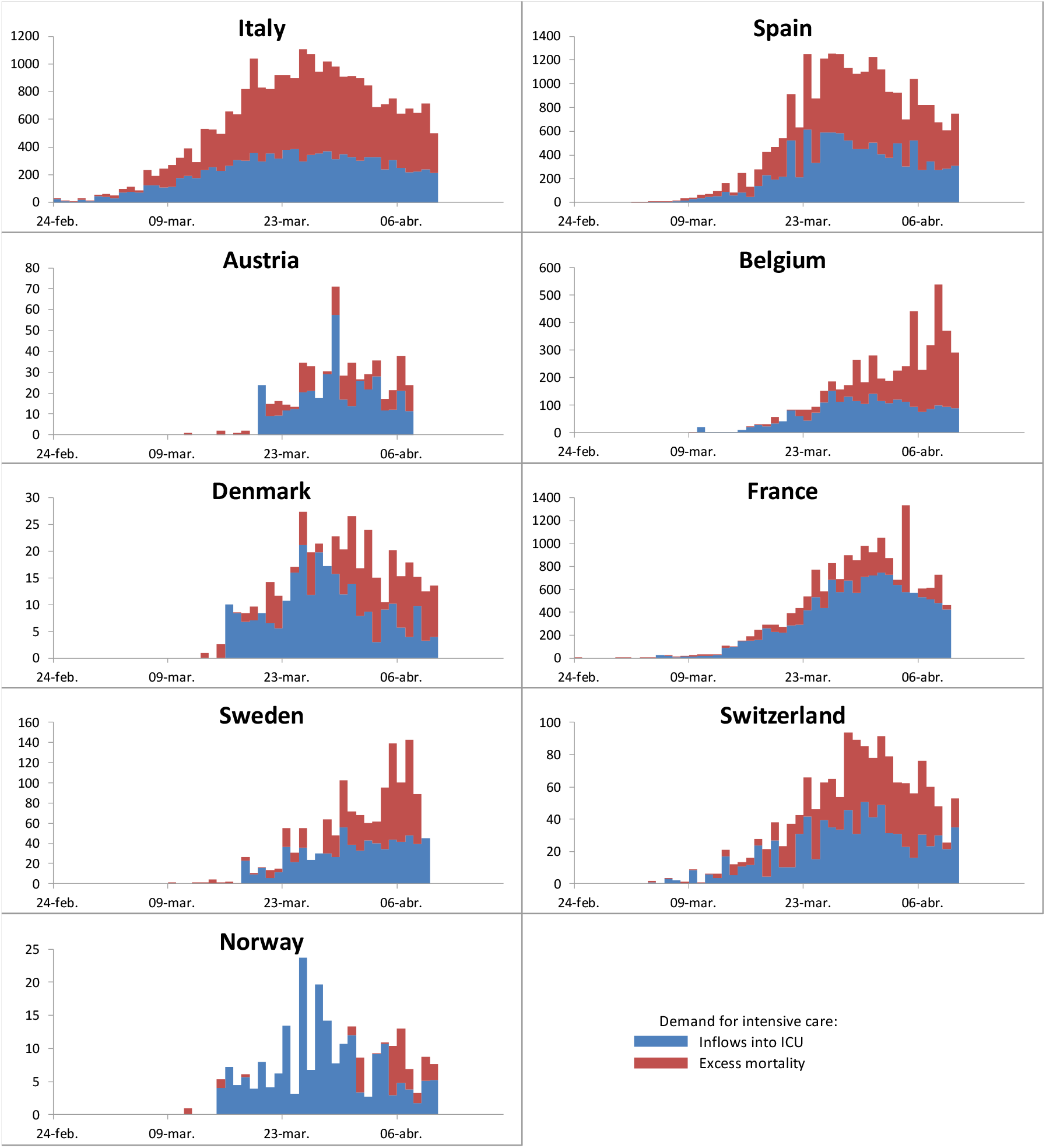
estimated demand for intensive care, as sum of inflows into ICU and excess mortality

## 3. Estimations of the effective reproductive number using ICU data

Now we plug the intensive care demand indicators presented in the previous section into the model of Flaxman et al (2020). Their codes are kept unchanged, including the database of nonpharmaceutical interventions implemented in each country, but our ICU indicator is used instead of the number of deaths (and the probability distribution that represents the time between the onset of symptoms and this event is reduced accordingly, i.e. we use a gamma distribution with a mean that is reduced by 7.5 days and coefficient of variation that is not altered).

The model estimates the effect of the nonpharmaceutical interventions that have been applied in these countries as a shift in the effective reproductive number that determines the rate of growth of the epidemic. Within the model, no other factors can affect R_t_ apart from these interventions.

Figure 3 presents the results of this time-varying estimation of R_t_ in each country. The first column replicates the original results with the original data (including also United Kingdom and Germany, even if they are not plotted in the figure). The second column updates the database and excludes United Kingdom and Germany, that don’t publish the necessary ICU data. And finally the third uses the model based on ICU demand.

**Figure 3:**
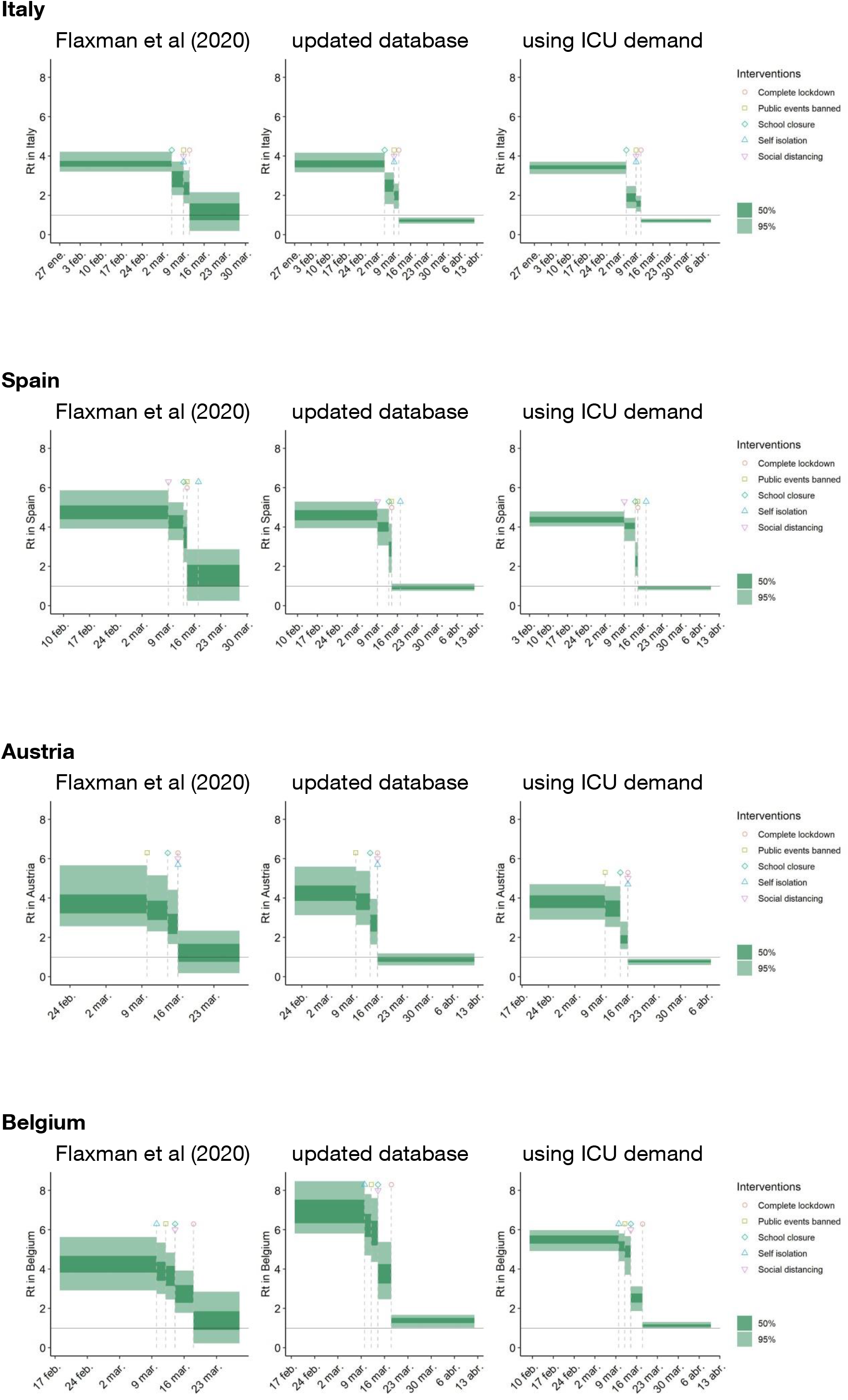
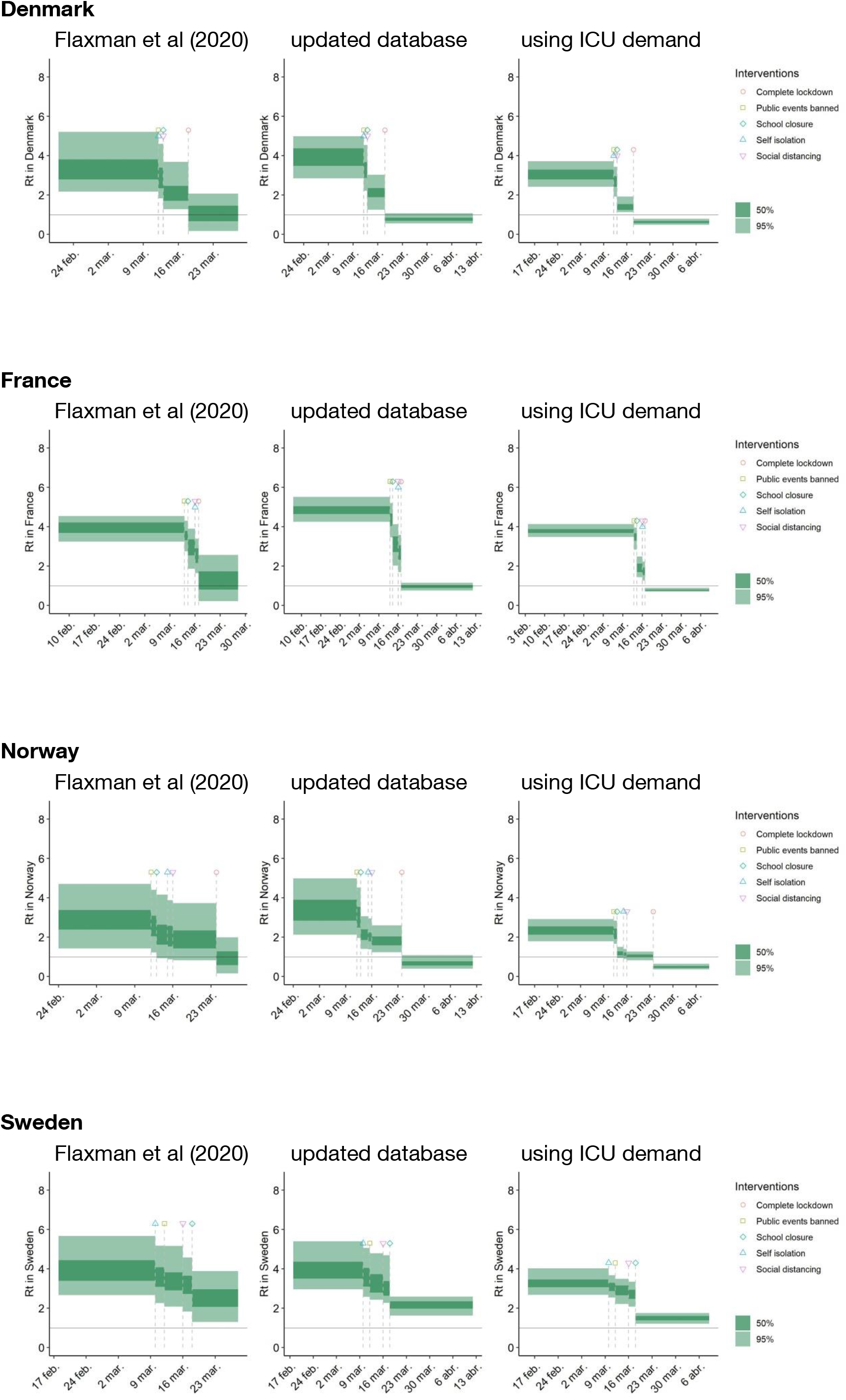
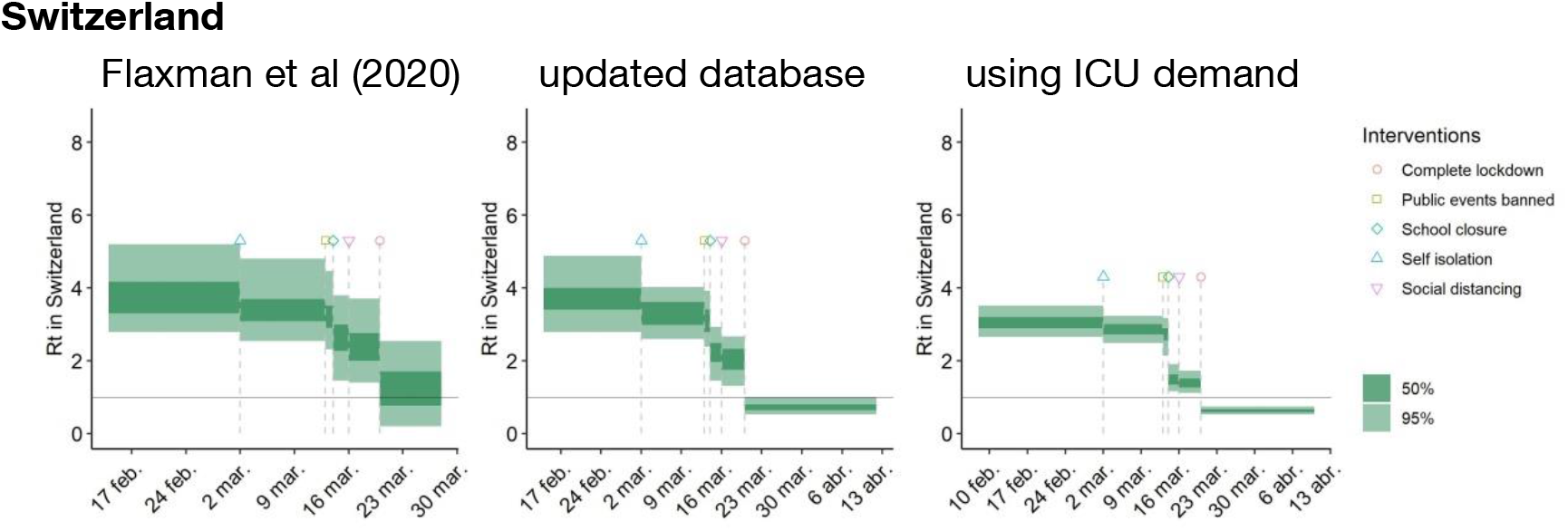
estimates of R_t_ and of the impact of nonpharmaceutical interventions following Flaxman et al (2020): with the original data, with an updated database as of 12^th^ of April, and adding the intensive care demand indicator presented in the previous section.

A result of R_t_ <1 with 95% confidence means that the interventions are enough to make the epidemic recede: instead of growing, it is becoming smaller every day. When Flaxman et al (2020) published their results, it was too early to conclude that this was the case in any of the countries considered. Updating the database with data available on the 12^th^ of April delivers an estimation of R_t_ <1 with 95% confidence in Italy and Switzerland, but not in the others: in most countries the estimation is inconclusive, with most of the mass of probability for R_t_ below 1 but some significant amount above 1 as well. **Using the intensive care demand indicator allows the estimation to conclude that we have reached R**_**t**_**<1 in 7 of the 9 countries considered: Italy, Spain, Austria, Denmark, France, Norway and Switzerland (but not Belgium or Sweden)**.

## Data Availability

All data used is publicly available: it is published by official institutions in each country.

https://github.com/daenuprobst/covid19-cases-switzerland

https://www.sst.dk/da/corona/tal-og-overvaagning

https://www.vg.no/spesial/2020/corona/

https://github.com/eschnou/covid19-be/blob/master/covid19-belgium.csv

https://www.mscbs.gob.es/profesionales/saludPublica/ccayes/alertasActual/nCov-China/documentos/Actualizacion_73_COVID-19.pdf

## Appendix: the ICU inflow-outflow model

Let *C*_*t*_ be the number of people in intensive care for COVID-19 at time *t, Ĉ*_*t*_ the cumulative number of people that have been in intensive care up to time *t*, and *C*_*it*_ and *C*_*ot*_ the inflows and outflows into/from intensive care at time *t*. Let *Ċ*_*i*t_ be the intensive care demand indicator that we want to calculate: the number of people who would need ICU admission at time *t*.

Let *D*_*t*_ be the number of COVID-19 patients who die at time *t*, and *Ďt* the estimation of the number of ICU patients that die at time *t*. Let *d*_*n*_ be the probability of dying *n* days after ICU admission, and *p*_*n*_ the probability of being discharged from ICU *n* days after admission (as presented in figure 1).

Then, for countries and regions that publish *C*_*t*_ (prevalence), we do the following: 

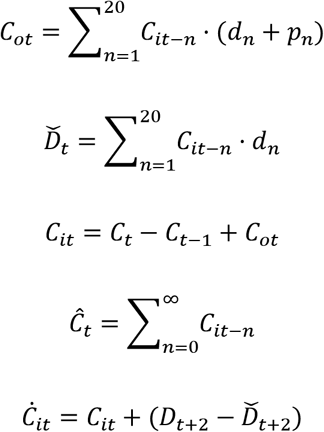

And for countries and regions that publish *Ĉ*_*t*_ (cumulative numbers), we do: 

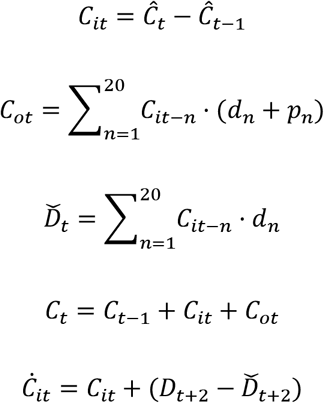

## Footnotes

1 Page 5 in Informe sobre la situación de COVID-19 en España, n° 21, 6 de abril de 2020, Ministerio de Sanidad, España.

## Notes

### Competing Interest Statement

The authors have declared no competing interest.

### Funding Statement

No external funding was received

